# Patient Trajectories from Electronic Health Records Suggest Specialty Specific Warning Signs for Common Variable Immunodeficiency

**DOI:** 10.64898/2026.10.01.26364525

**Authors:** Rachel Mester, Aaron T. Chin, Veronica Tozzo, Alexis V. Stephens, Hal M. Hoffman, Marc A. Reidl, Morna J. Dorsey, Sudhir Gupta, Yesim Y. Demirdag, Sriram Sankararaman, Lisa Bastarache, Bogdan Pasaniuc, Manish J. Butte

## Abstract

**Background:** Common variable immunodeficiency is frequently underrecognized due to its heterogeneous clinical presentation, resulting in diagnostic delays that increase disease-related complications, with each year of delay increasing mortality risk by 4% for the most common inborn error of immunity.

**Objective:** In individuals with established common variable immunodeficiency, we sought to recognize patterns in patient diagnostic trajectories to identify opportunities for interventions to reduce time to diagnosis.

**Methods:** We analyzed retrospective electronic health records data from 139 individuals with physician-confirmed common variable immunodeficiency across five academic health systems representing >10million patients. We evaluated diagnosing specialties and international classification of disease code trajectories in the two years prior to and at common variable immunodeficiency confirmation. Additionally, we construct specialty-specific prediction on a 1:50 case: control cohort using a gradient boosting algorithm.

**Results:** The leading diagnoses at common variable immunodeficiency determination were respiratory (35% of individuals), and individuals with respiratory symptoms were more likely to be diagnosed by Allergy / Immunology than by other specialties (odds ratio 2.9, standard error 0.35, p-value 1.9x10^-3^) and had a higher frequency of visits to pulmonary disease in the two years prior to diagnosis. Individuals with neoplasms were 50% more likely than other patients (OR 2.0, SE 0.29, p-value 0.02) to receive a diagnosis at each visit, reducing their diagnostic delay by up to a year. We find that a larger ratio of number of specialties visited to the total number of visits is significantly correlated with an increased time to diagnosis. A trajectory-informed specialty-specific prediction improved detection with an area under the precision-recall curve of fifty times the value expected under the null.

**Conclusions:** Individuals referred to Allergy / Immunology have shorter time to diagnosis, and a specialty-specific prediction could improve detection of common variable immunodeficiency.

**Clinical Implications:** Overall, we find that phenotype-specialty interactions may inform targeted interventions to improve common variable immunodeficiency recognition across specialties, reducing diagnostic odyssey and disease-related complications.

**Capsule Summary:** Specialty-specific symptom patterns and patient trajectories preceding common variable immunodeficiency diagnosis reveal opportunities for earlier recognition through context-aware electronic health record–based prediction.

## Introduction

Common Variable Immunodeficiency (CVID) encompasses a group of conditions characterized by impaired antibody production and represents the most common symptomatic antibody deficiency diagnosed in adults, estimated to impact up to 300,000 individuals worldwide.^1^ Symptoms typically emerge between the second and fourth decades of life, with significant phenotypic variability complicating diagnosis.^2,3^ Many patients present with recurrent infections, but a substantial proportion present with noninfectious immune dysregulation.

Timely diagnosis for individuals with CVID is important. For each year a diagnosis is delayed, there is an estimated 4% increase in mortality.^3–6^ However, diagnostic delay is common, with previous studies measuring mean diagnostic delay at up to 10 years, with noninfectious complications representing the biggest challenge to reducing mortality.^7^ Clinical recognition of CVID is difficult because presenting symptoms are highly heterogeneous and can mimic common conditions.

Compared to those unaffected, individuals with CVID have more clinic visits in the years leading up to their diagnosis, including more consultations with their general practitioners than similar individuals without CVID.^8^ Even with an expert, one study found that it took an average of seven outpatient visits or one inpatient admission to achieve the correct diagnosis.^9^ A previous study analyzing over 78,000 primary care visits found a strong inverse relationship between the prevalence of a condition at each practice and the rate at which individuals with the conditions are referred out of that practice (r=-0.87).^10^ Other factors such as age at symptom onset and socioeconomic status, alter the path to a CVID diagnosis.^11,12^ Despite public health campaigns aimed at increasing awareness of primary immunodeficiencies, studies reveal persistent gaps in physician knowledge.^13–15^ Even targeted educational programs have struggled to correct misconceptions, such as the belief that CVID is primarily diagnosed in children.^16^

Understanding the diagnostic journey taken by individuals with CVID is crucial to identifying targeted interventions that could reduce diagnostic delays and associated complications.^15^ But the exact nature of these diagnostic journeys is unclear. Understanding where undiagnosed CVID patients receive care and the relationship between care context and presentation could improve interventions meant to reduce diagnostic delay, such as educational initiatives or the implementation of patient finding algorithms.

In this study, we leverage electronic health record (EHR) data from a diverse multi-institutional cohort of individuals with established CVID to map referral and phenotypic patterns to offer a real-world view of the diagnostic journey. We examine which specialties individuals with CVID visit prior to diagnosis, what symptoms are present at their diagnostic visit, and test the hypothesis that clinical presentation differs across clinical context.

## Methods

### Cohort Definition

Building on our previous study,^17^ we considered a case cohort of 558 individuals with CVID whose diagnosis was ascertained by clinical immunologists at five University of California (UC) health systems - Los Angeles, San Francisco, Irvine, San Diego, and Davis. These individuals ranged in age from 11 years (y) to 89y. Data was accessed through the UC Health Data Warehouse, a de-identified database with 8.7 million patients following the Observational Medical Outcomes Partnership data standard.^18,19^

Starting with this case cohort of 558 individuals, we then selected patients into the trajectory cohort for this study based on criteria to ensure sufficient longitudinal EHR history to diagnosis. See eMethods for full eligibility criteria (Figure E1). The resulting cohort consisted of 139 individuals with the same age range as the original cohort, and with a mean pre-diagnosis immunoglobulin G (IgG) of 425 milligrams per deciliter (mg/dL) (Table EII).

### Phecode-Based Clinical Phenotyping

Patient symptoms and diagnoses were captured in the UC Health Data Warehouse as international classification of disease (ICD) 9/10 codes linked to individual encounters. Only inpatient and outpatient visits were included. We used phecodes, which are groupings of ICD-9/ICD-10 diagnosis codes into clinically meaningful phenotypes.^20^ Phecodes were further consolidated by merging all phecodes sharing a pre-decimal code, reducing granularity to broader clinical entities (e.g. 474.1 to 474). For analyses requiring higher-level groupings, phecodes were categorized based on the PheWAS catalog (e.g. chronic sinusitis to respiratory).^21^ Phecodes determined to be clinically irrelevant to CVID, including codes reflecting acute, incidental, or unrelated conditions, were excluded from all eligibility criteria and downstream analyses. The full exclusion list is provided in Table EI.

### Diagnostic Specialty and Diagnosis Determination

We determined the specialist who initially diagnosed the patient (diagnostic specialty) by linking the visit of the index encounter to the provider’s department or, in absence of a specified department or provider, the specialty of the care site. When the date of diagnosis corresponded to a lab encounter, we considered the specialty of the chronologically preceding visit. We considered diagnoses as recorded by ICD 9/10 codes at inpatient and outpatient visits; problem list, historical, and billing-only diagnoses were not included.

### Finding Matched Controls

For each case, we identified 50 controls with the same age, sex, self-reported race, and health system, and similar EHR length (within 180 days). Controls were required to have at least one visit to the specialty in which their matched case was diagnosed.

### Time to Diagnosis Analysis

We estimated the associations between presence of specific CVID-related ICD codes and time to CVID diagnosis using Cox proportional hazards regression models fit with the lifelines CoxPHFitter function.^22^ Follow-up time was defined as the interval between an individual’s first visit with a CVID-related phecode and the index encounter. Because all individuals had an index encounter, no censoring was required. Kaplan-Meier plots were used to visualize time to diagnosis across phenotype groups and the “bounce” score, which we defined as the ratio of the number of specialties visited to the total number of visits (see eMethods).

### Specialty-Specific CVID Prediction

For our specialty-specific CVID prediction analysis, we trained a gradient-boosted decision tree model using XGBoost to differentiate between CVID cases and the matched controls, specific to the specialty at which each CVID patient was diagnosed. The data for each specialty was split into a training (80%) and testing (20%) split, used for model training and evaluation. We then calculated area under the receiver-operator curve (AUROC) and area under the precision-recall curve (AU-PR), as well as the Shapley values of the features in the model using the TreeExplainer module in the shap python library.^23^

## Results

### Diagnosing specialty of CVID varies by respiratory symptom status

We stratified our 139 patient cohort of patients by age (<26y, 26-64y, >64y) and the specialty at which they received their CVID diagnosis (Figure 1). Of the six patients <26y, four had a diagnosis specialty of Pediatric Medicine, considered to be part of General Practice. For the 35 patients 26-64y and the 53 patients >64y with an identifiable diagnosis specialty, only 26% were diagnosed in any department considered General Practice. Allergy / Immunology diagnosed 11 patients 26-64y and 18 for patients >64y. Rheumatology was one of the most common diagnosing specialties for patients 26-64y, while Gastroenterology and Hematology were among the most common diagnosing specialties for patients over 64y. Pulmonary Disease was the most common diagnosing specialty for all patients over 25y. Thus, we find that while General practice and Allergy / Immunology do account for a large proportion of CVID diagnosis, other specialties, notably Rheumatology, Pulmonary Disease, and Gastroenterology, also played a role in diagnosis in our cohort.

**Figure 1:**
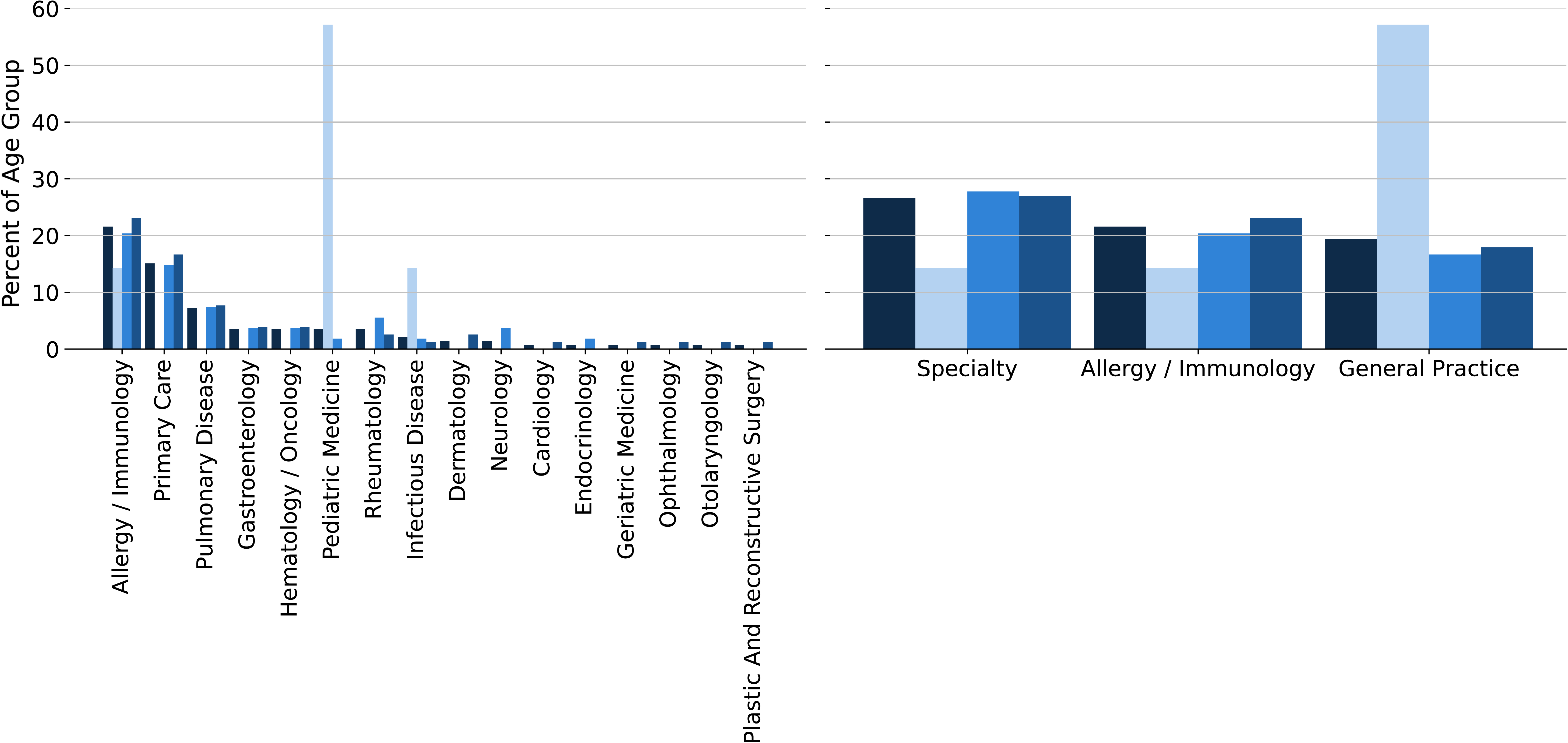
This figure shows the clinical specialty of patients’ CVID diagnosis visits (x axis). Colors indicate percentages of different age groups. The left plot displays the specific specialties present in our cohort at the diagnosis visit. The right plot displays these specialties grouped by category.

Respiratory ailments account for a large proportion of CVID symptoms in our patient cohort at the time of diagnosis. When we also stratified our cohort by phecode category present at individuals’ index encounters (Figure E2), we found that respiratory symptoms were the most common type of symptom, with 40% of individuals >64y receiving a respiratory diagnosis during their CVID diagnosis visit. While we previously noted that many patients were diagnosed in pulmonary disease specialty clinics, many individuals not diagnosed by pulmonary disease also received respiratory diagnoses during their CVID diagnosis visit.

We compared the odds of an individual being diagnosed at an Allergy / Immunology clinic having various CVID symptoms compared to individuals diagnosed at any other specialty having those symptoms (see eMethods). The goal of this analysis was to understand if there are vital differences to patients who were referred to Allergy / Immunology before versus after CVID diagnosis, and if those differences could be used to help future patients receive referrals to Allergy / Immunology sooner. We found that individuals with respiratory symptoms had a higher odds of being diagnosed in Allergy / Immunology clinics, specifically with an odds ratio (OR) as high as 2.9 with standard error (SE) 0.35 and p-value 1.9x10^-3^ chronic sinusitis (Figure 2). Individuals with other infections were also more likely to be diagnosed at Allergy / Immunology than other departments, with OR 11.0 (SE 1.1, p-value 0.03). By contrast, some OR, although not significant due to sample size, were suggestive that individuals would be less likely to be diagnosed in an Allergy / Immunology clinic. Individuals with gastrointestinal symptoms were only half as likely to be diagnosed in an Allergy / Immunology clinic versus elsewhere (OR 0.49), SE 0.67, p-value 0.28). Similarly, individuals with blood disorders had OR 0.66 (SE 0.60, p-value 0.49), a finding that is driven by the large numbers of the older population in our cohort having been diagnosed in a Hematology clinic.

**Figure 2:**
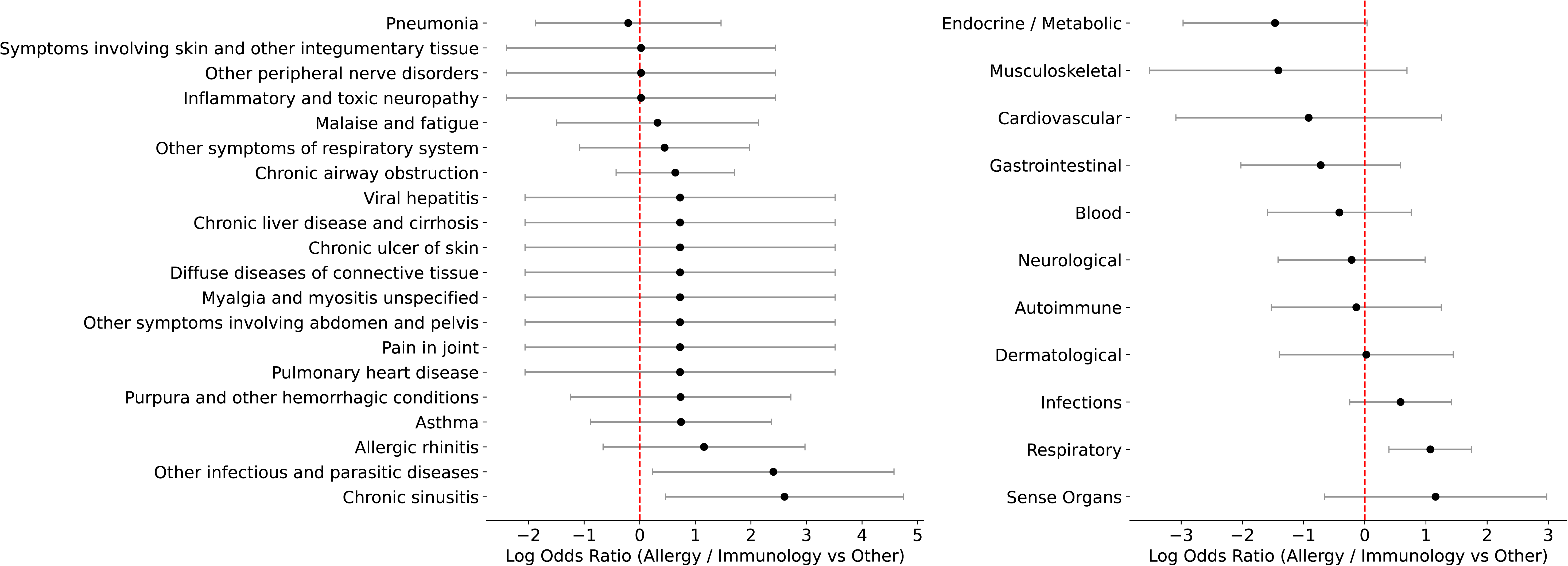
This figure shows the log odds ratios and 95% confidence intervals of patients diagnosed at Allergy / Immunology compared to other specialties depending on the phecodes (left) or phecode categories (right) present at their diagnosis visit.

### CVID patients bounce between specialties prior to diagnosis

After collecting data on our cohorts’ health care visit history over the two years prior to CVID diagnosis (see eMethods), we found that some individuals with CVID had up to 30 visits with Primary Care during the two years prior to diagnosis (Figure E3). This result reflects the heightened healthcare utilization that precedes CVID diagnosis for most individuals. Some individuals in the >64y cohort had high numbers of visits to pulmonary clinics, reflecting the high prevalence of pulmonary disease diagnosis among those individuals. For subjects aged 26-64y, visits to gastroenterology clinics were more common than any other specialty aside from Allergy / Immunology. More obvious than any specific pattern was the finding that everyone in our cohort has a unique fingerprint of visits during this time, highlighting the heterogeneity of CVID disease trajectory.

To visualize the order in which patients were referred to various specialties prior to CVID diagnosis, we mapped individuals’ trajectories using a network plot, excluding individuals <26y due to small sample size (Figure 3). For both age groups shown, we saw that the most likely outcome of a visit to Allergy / Immunology was a CVID diagnosis (89% in 26y-64y and 79% in >64y). By contrast, individuals seen by a gastroenterology clinic were more likely to revisit gastroenterology clinics multiple times before a CVID diagnosis was made (88% in 26y-64y and 38% in >64y). Individuals seen by pulmonary disease clinic were more likely to receive a CVID diagnosis in the 26-64y age group (46%), and less likely in the >64y age group (11%). These findings indicate a potentially age-specific interaction between specialty clinics and CVID diagnosis, in which younger individuals visiting a pulmonary clinic are considered more likely candidates for CVID than their older counterparts.

**Figure 3:**
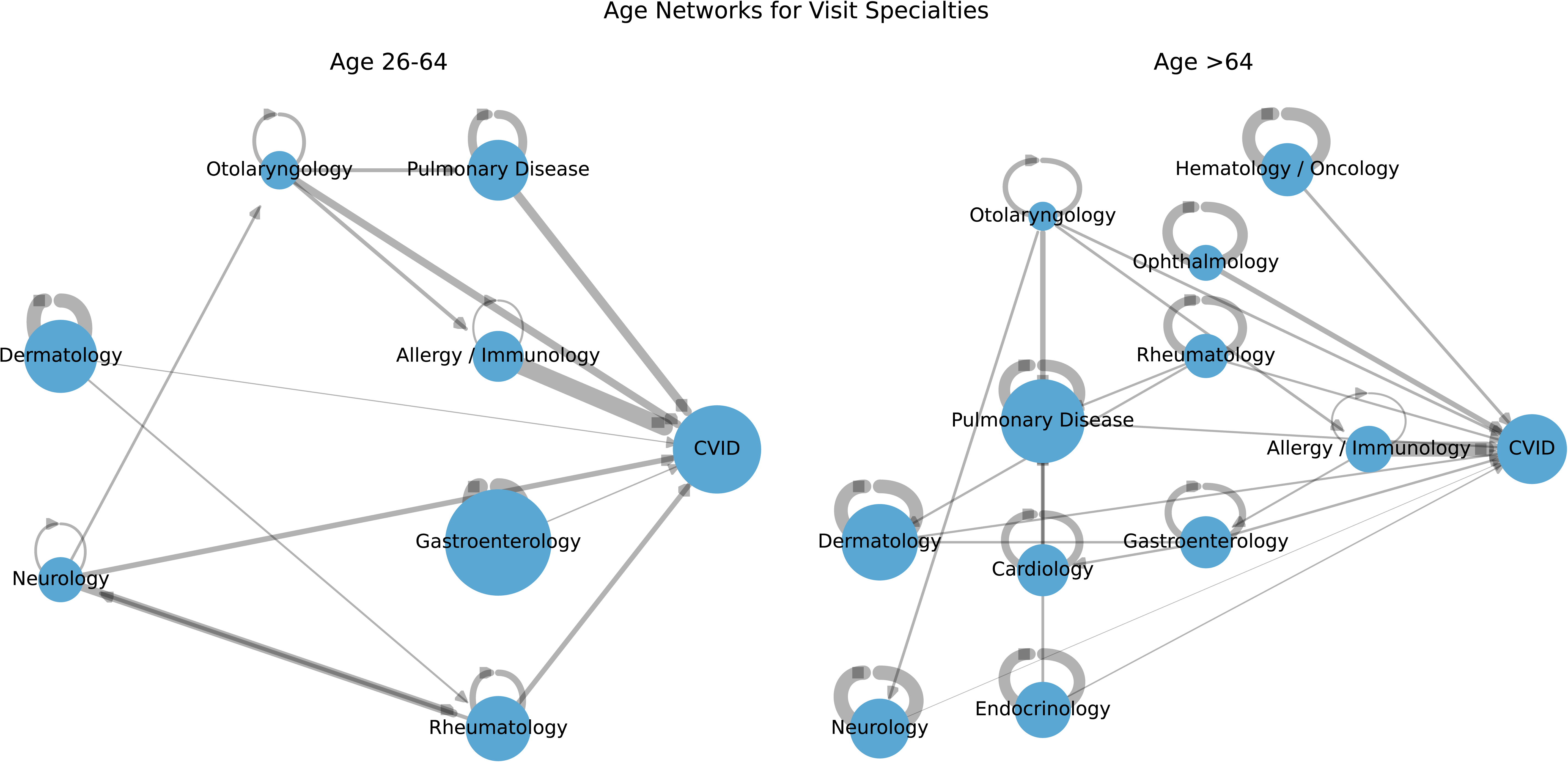
Patients 26-64y (left) and >64y (right) have distinct trajectories through various clinical specialties prior to CVID diagnosis. Nodes represent specialties; edges represent subsequent visits. Circle size indicates specialty prevalence, and thicker arrows indicate a higher probability that a patient’s next visit is to the specialty across the arrow. Each patient’s journey ends with CVID diagnosis.

To more properly understand if providers are seeing individuals with CVID more frequently than other individuals, we compared our cohort with 6,950 matched controls (see Methods). We compared the number of visits to the diagnosing specialty and total specialties seen during the two years prior to diagnosis (Figure E4). We found that, apart from Pediatric Medicine, individuals with CVID were not visiting the specialty at which they received their CVID diagnosis significantly more times in the two years prior to diagnosis than the average matched control of that specialty (p-value 0.09, Kolmogorov-Smirnov test). However, we saw that individuals diagnosed in a pulmonary clinic showed a bimodal distribution, where some (but not all) individuals with CVID experienced getting “stuck” in pulmonary clinic(s) for many repeated visits prior to receiving a CVID diagnosis. We also saw that for all individuals under 65, those with CVID saw a higher number of different specialty clinics in the two years prior to diagnosis compared to controls receiving similar care. This finding reveals that for the most part, individuals with CVID do not visit specific specialties more often than matched controls from the same specialty. They do however visit more different specialties than those same controls, indicating that diagnostic delay is more likely to result from bouncing around between specialties as opposed to getting stuck in any one specialty.

### Neoplasms decrease time to diagnosis for individuals with CVID

To investigate whether any diagnosis occurring in the two years prior to diagnosis impacted time to diagnosis, we estimated the hazard ratios using symptom categories as multivariable exposures (Figure 4). The model also included a “bounce” score (see Methods), which indicated how many different specialties a patient saw. The mean (SD) score was 0.57 (0.31) in our cohort. The Cox model trained had a concordance of 0.76 and a p-value <6x10^-10^, indicating that it could discriminate between individuals with a generally higher risk of diagnostic delay and those without. We found that individuals with neoplasms, neurological, or musculoskeletal diagnoses received a CVID diagnosis faster than individuals without these symptoms, to a nominally significant degree. This difference can potentially be attributed to the more extensive testing undergone by these patient populations. The most significant feature was the “bounce” score. Individuals with a higher-than-average bounce score were 93% less likely to receive a CVID diagnosis at each visit than individuals with lower-than-average bounce scores. Because the bounce score automatically for the total number of visits for each patient, this result suggests that bouncing between different specialties, rather than a sheerly large number of visits, accounts for a significant portion of diagnostic delay.

**Figure 4:**
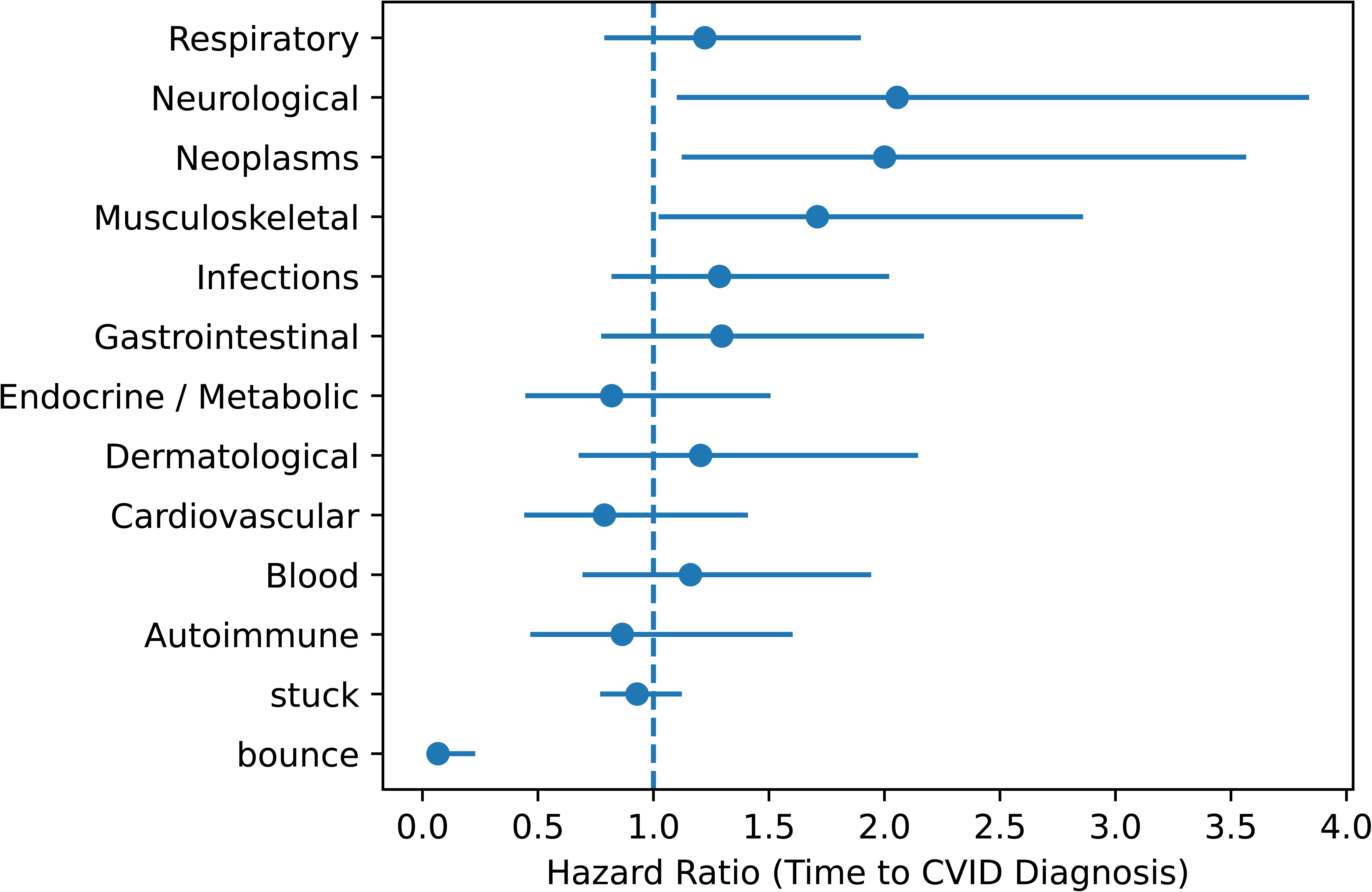
Forest plot for time to CVID diagnosis. The x axis shows the hazard ratio for a patient receiving a CVID diagnosis with and without the feature noted on the y-axis. A hazard ratio of 1 indicates no difference, <1 indicates that the feature prolongs the time to diagnosis, and >1 indicates a decrease in time to diagnosis.

We visualized the differences in diagnosis time for two of the most significant features, bounce score and neoplasms (Figure 5). Over 80% of individuals with high bounce scores had over a year-long diagnostic journey compared to only about 40% of low bounce individuals. On the other hand, only 30% of individuals with neoplasms had a documented diagnostic journey of longer than one year, compared to over 75% of those without.

**Figure 5:**
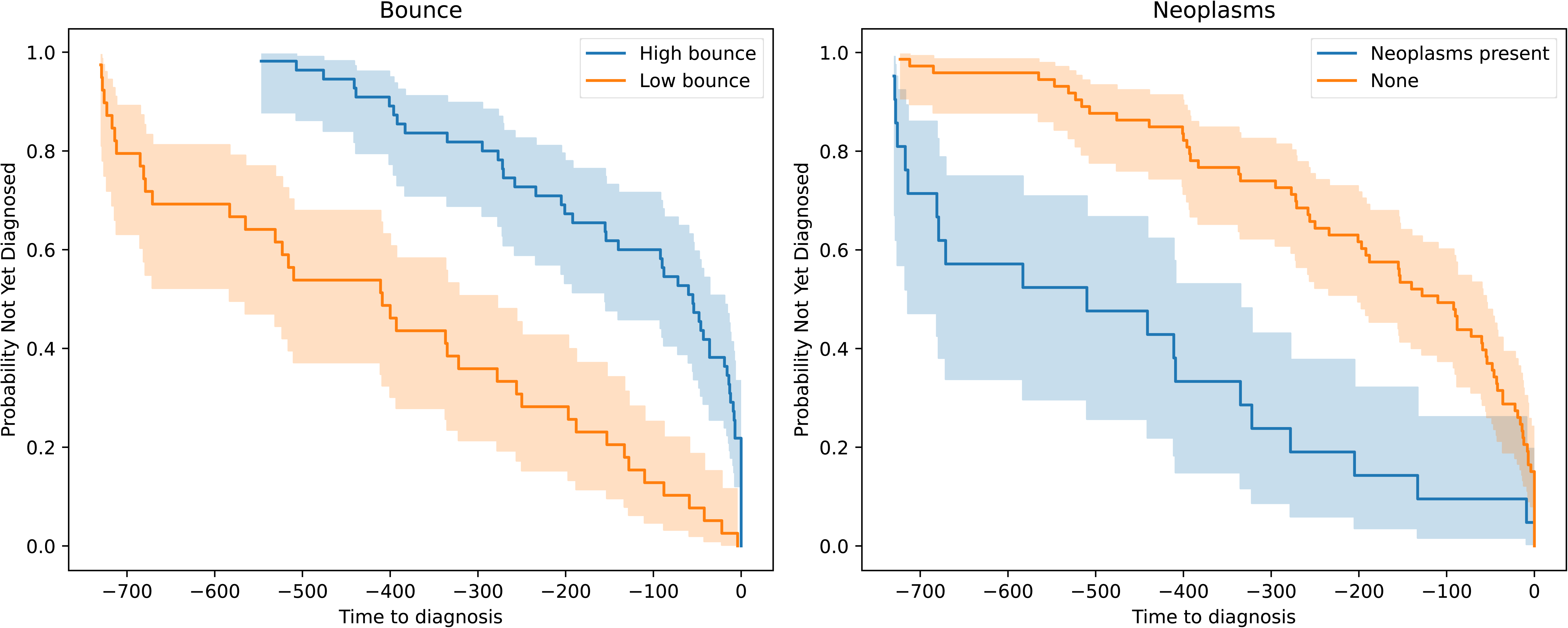
Stratified Kaplan-Meier curves for time to CVID diagnosis. Left: CVID patients with above (blue) or below (orange) median bounce score. Right: CVID patients with (blue) and without (orange) neoplasms. Both: The x-axis displays the time prior to diagnosis. The y-axis indicates the probability that a patient has not yet been diagnosed.

Considering that many patients did not have a long EHR history, we also considered whether the length of our “lookback window” impacted how these features shaped time to diagnosis (Figure E5). Patients with neoplasms within 6 months of CVID diagnosis were more likely to have had a shorter diagnostic journey, whereas neoplasms two years prior to diagnosis did not have the same effect. This indicates that the presence of neoplasms was an important factor that led to rapid referral diagnosis of CVID and is driven by the individuals diagnosed with CVID in hematology / oncology clinics.

### Identifying specialty-specific predictors of CVID diagnosis

We further examined whether clinical phenotypes associated with CVID differed across medical specialties using specialty-matched case–control prediction. The primary purpose of this analysis was to determine if there were specialty specific indicators that the diagnosing providers had utilized in order to make their diagnoses that we could observe in the data as a pattern and suggest as specialty-specific warning signs to be used more broadly by providers of that specialty. We did observe distinct and reproducible phenotypic signatures across specialties (Figure 6). In analyzing which factors drove the time to diagnosis across each specialty clinic, the neoplasm signal was most prevalent in patients diagnosed in Allergy / Immunology or pulmonary disease clinics. Furthermore, CVID patients had statistically significant higher odds of having neoplasms than other patients of pulmonary disease. We found that CVID cases demonstrated enrichment of multiple symptom categories relative to controls, but the magnitude and pattern of associations varied substantially by specialty. Other phenotypes also appeared more specialty-specific, suggesting heterogeneity in clinical presentation and diagnostic pathways. For example, patients presenting in a pulmonary clinic with gastrointestinal symptoms were indicative of CVID, whereas patients presenting in a gastroenterology clinic with those same symptoms were not. These results highlight that CVID prediction depends on the clinical context in which patients are evaluated.

**Figure 6:**
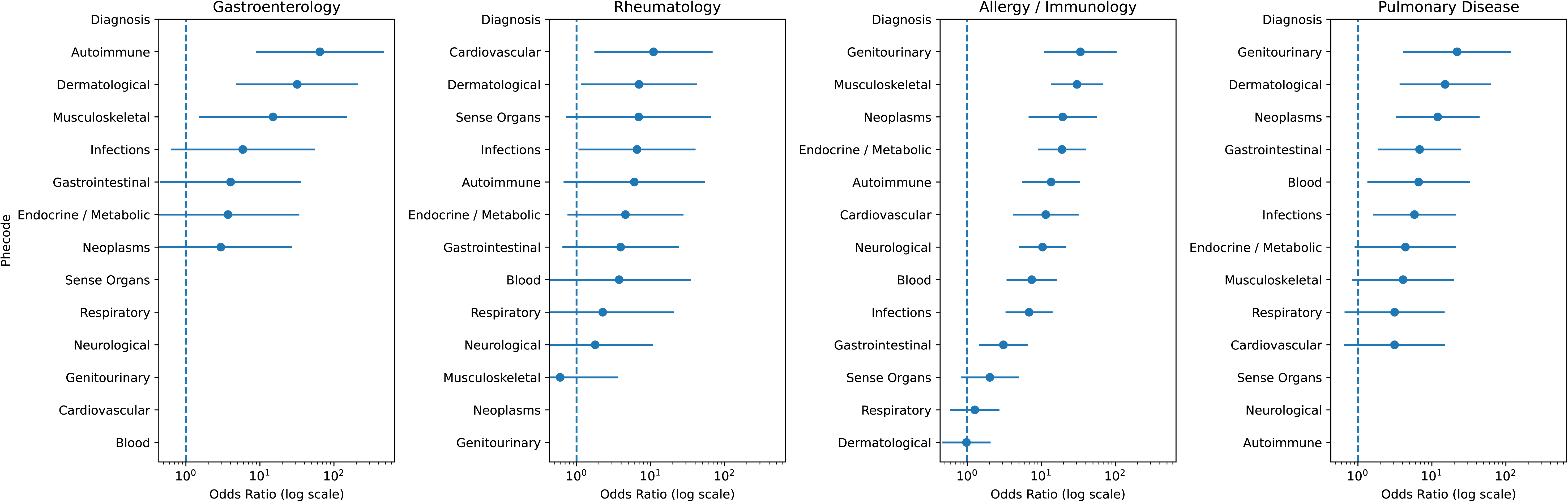
This figure shows the odds ratios for each type of symptom for patients diagnosed in each of these specialties. The specialty of diagnosis is the panel, and the phecode categories are represented by rows. The odds ratios (x-axis) compare CVID cases diagnosed in that specialty with matched controls seen by that specialty.

To evaluate whether these heterogeneous phenotypic signatures could be leveraged for prediction, we trained specialty-specific models to classify CVID cases versus controls (Figure E7). Across specialties, the models demonstrated variable but generally strong discrimination, with AU-ROC of 0.90 in gastroenterology and 0.94 in pulmonary disease compared to an expectation of 0.5 under a null model. Precision-recall (PR) curves further highlighted meaningful performance in the context of class imbalance, with AUC-PR consistently of 0.76 for gastroenterology and 0.51 for pulmonary disease, compared to an expected AUC-PR of 0.02 under a null model. Even after bootstrapping results over five iterations, the AUC-PR and AUC-ROC were 0.53 and 0.79 respectively for gastroenterology and 0.52 and 0.93 respectively for pulmonary disease. These findings show that predictive signals for CVID are present in the clinics in which CVID patients are presenting, providing opportunity for earlier diagnosis and reduced diagnostic delay.

To identify the clinical features driving predictions, we used SHAP (Shapley Additive Explanations) to interpret the specialty-specific XGBoost models (Figure E6). Each specialty exhibited a distinct hierarchy of predictive features, reflecting differences in the symptom profiles most informative for CVID classification across clinical contexts. For example, the biggest drivers of determining CVID status for patients in gastroenterology are autoimmune, musculoskeletal, and dermatological symptoms. Meanwhile the largest drivers of determining CVID status for patients in pulmonary disease clinics are gastrointestinal, dermatological, and neoplastic symptoms. Collectively, these results demonstrate that while there is overlap in key predictive features, each specialty leverages a partially distinct clinical signature to identify patients at risk of CVID. For example, dermatological symptoms are associated with CVID status for both pulmonary disease and gastrointestinal clinics, indicating that for patients seen in these specialties, dermatological symptoms are uncommon unless the patient is suffering from a systemic illness such as CVID. However, gastrointestinal symptoms are associated with CVID status when they are addressed in a pulmonary disease clinic, but not when addressed in gastroenterology.

## Discussion

In this multi–health system aggregated analysis, we demonstrate that the diagnostic journey for CVID is strongly shaped by specialty-specific phenotypic patterns and healthcare utilization trajectories. For example, individuals diagnosed in Allergy/Immunology were more likely to present with respiratory and infectious phenotypes, consistent with established clinical teaching that recurrent sinopulmonary infections are a hallmark trigger for immunologic evaluation. Another example is that individuals who were seen in a pulmonary clinic for pulmonary symptoms, but who also had dermatological conditions were much more likely to eventually be diagnosed with CVID than those presenting with pulmonary symptoms alone.

We show that individuals under 65y with CVID engage in broader healthcare utilization than matched controls in the years preceding diagnosis, consistent with prior reports of increased healthcare contact in undiagnosed CVID^4^. We demonstrate that individuals with CVID systematically traverse multiple specialties prior to diagnosis, indicating that diagnostic delay is driven in part by cross-specialty fragmentation rather than within-specialty “tunnel vision”.

Time-to-diagnoses analyses further reveal phenotypes that were correlated with accelerated or delayed recognition. For example, neoplastic diagnoses were associated with shorter time to CVID diagnosis. Neoplasms often trigger broader systemic evaluation that increases the likelihood of immunologic workup. Specifically, patients with malignancies were 5 times more likely to be referred out of primary care than those with respiratory illnesses.^10^ In contrast, the ratio of number of specialties visited to the total number of visits (bounce score) was strongly associated with prolonged time to diagnosis. Individuals with higher bounce scores experienced substantially delayed diagnosis, consistent with a model in which heterogeneous symptoms lead to diffuse evaluations without diagnostic convergence. This is a particularly useful finding because not only is a high bounce score a predictor of diagnostic delay, but it is also shown to be higher in individuals with CVID than matched controls, providing an opportunity for intervention that is identifiable in real time within the EHR.

We further demonstrate that specialty-specific predictive models can leverage these heterogeneous phenotypic patterns to distinguish CVID cases from matched controls in our cohort. While our cohort is too small to expect these specific scores to translate outside of our dataset, our success with this data reveals that specialty-specific prediction algorithms have promise in the objective of reducing CVID diagnostic delay. Heterogeneity in feature importance across specialties underscores that CVID prediction is not driven by a universal set of features, but rather by context-dependent combinations of symptoms and utilization patterns.

These results have practical implications for intervention design. First, they support the development of specialty-specific clinical decision support tools that identify high-risk patients within existing workflows. Second, they suggest that referral pathways represent a key leverage point for reducing diagnostic delay. Third, they indicate that trajectory-based features, such as cross-specialty dispersion, may provide actionable signals for identifying individuals who warrant earlier immunologic evaluation.

This study has several limitations. Our cohort had a small sample size and represented a subset of individuals with sufficient longitudinal documentation. Findings may therefore not generalize to the general population. Matched controls, while used to approximate specialty specific clinical populations, cannot fully eliminate residual confounding by healthcare access and comorbidity burden. Finally, our analyses are restricted to pre-diagnostic trajectories and do not capture post-diagnostic outcomes or treatment effects.

The path to CVID diagnosis is shaped not by symptoms alone, but by specialty context that frames how those symptoms are interpreted. Specialty-specific warning signs offer concrete, EHR derived indicators that could be operationalized within clinical decision support systems to flag individuals warranting earlier immunologic evaluation. Prospective validation of these signals represents a clear next step toward translating this trajectory-based framework into earlier CVID recognition and reduced diagnostic morbidity.

## Supporting information

eMethods

## Data Availability

All data produced in the present study are available upon reasonable request to the authors.

## Acknowledgements

This research made possible by funding from the National Institutes of Health (NIH) / National Institutes of Allergy and Infectious Diseases R01 AI153827 to M.J.B. and B.P. The authors thank the Center for Data-driven Insights and Innovation at UC Health for its analytical and technical support related to use of the UC Health Data Warehouse and related data assets.

## Abbreviations

CVID: common variable immunodeficiency
EHR: electronic health record
UC: University of California
ICD: international classification of diseases
IgG: immunoglobulin G
mg/dL: milligrams per deciliter
SD: standard deviation
AUROC: area under the receiver operator curve
AU-PR: area under the precision-recall curve
y: years
OR: odds ratio
SE: standard error

