## Supplementary material for "Patient Trajectories from Electronic Health Records Suggest Specialty Specific Warning Signs for Common Variable Immunodeficiency": eMethods

BOX 951766, Room 5303 Life Sciences

Los Angeles, CA 90095-1766

### **eMethods**

*Patient Trajectory Cohort Eligibility Criteria*

A patient’s index encounter was determined to be the patient’s first instance of a CVID ICD code (279.06 / D83*) or an IgG lab result of less than 600 mg/dL. Patients were then required to have at least 45 days of EHR history prior to the index encounter. Patients were required to have at least one phecode in the two years prior to their index encounter. Phecodes determined to be clinically irrelevant to CVID were excluded from this list and the subsequent analyses (Table EI). Any patient with an IgG lab result of >600 milligrams per deciliter (mg/dL) prior to CVID diagnosis was excluded from subsequent analyses due to the likelihood that the patient had been previously diagnosed and begun receiving immunoglobulin replacement therapy elsewhere prior to coming to the UC.

*Bounce Score Definition*

The bounce score was defined as: (Number of distinct specialties visited) / (Total number of visits)

### A higher bounce score reflects care distributed across many specialties relative to overall visit volume, consistent with a diagnostic odyssey in which patients are evaluated across departments without a unifying diagnosis. The score was computed over each patient's pre-index encounter period.

*Patient Journey Visit Tabulation*

To count individual visits to each specialty in the two years prior to CVID diagnosis, we accumulated all visits that led up to every patient in our cohort receiving their CVID diagnosis. We used the same phecode filters that we used to determine patient EHR history of relevant phecodes, and the same method of determining the visit specialty as we did for the diagnostic specialties.

*Association Analysis of CVID Symptoms and Allergy / Immunology Diagnosis*

To compare CVID symptoms at diagnosis between Allergy / Immunology and other specialties, we collapsed patient symptoms into binary variables (1 if they had the symptom at diagnosis and 0 if they hadn’t). We then computed he odds ratio that a patient who had a particular symptom was diagnosed at Allergy Immunology.

### Table EI: Phecode Exclusions

| **Phecode** | **Phenotype** |
| --- | --- |
| 174 | breast cancer |
| 175 | acquired absence of breast |
| 225 | benign neoplasm of brain and other parts of nervous system |
| 276 | disorders of fluid, electrolyte, and acid-base imbalance |
| 278 | overweight, obesity, and other hyperalimentation |
| 291 | other unspecified nonpsychotic and / or transient mental disorders |
| 295 | schizophrenia and other psychotic disorders |
| 296 | mood disorders |
| 297 | suicidal ideation |
| 300 | anxiety, phobic and dissociative disorders |
| 305 | eating disorder |
| 306 | other mental disorder |
| 313 | pervasive developmental disorders |
| 316 | substance addiction and disorders |
| 318 | tobacco use disorder |
| 327 | sleep disorders |
| 340 | migraine |
| 350 | abnormal movement |
| 355 | complex regional / central pain syndrome |
| 361 | retinal detachments and defects |
| 365 | glaucoma |
| 366 | cataract |
| 379 | other disorders of the eye |
| 386 | vertiginous syndromes and other disorders of vestibular system |
| 395 | heart valve disorders |
| 401 | hypertension |
| 411 | ischemic heart disease |
| 427 | cardiac dysrhythmias |
| 429 | ill-defined descriptions and complications of heart disease |
| 443 | peripheral vascular disease |
| 453 | chronic venous hypertension |
| 454 | varicose veins |
| 500 | lung disease due to external agents |
| 525 | other diseases of teeth and supporting structures |
| 539 | bariatric surgery |
| 550 | abdominal hernia |
| 594 | urinary calculus |
| 599 | other symptoms / disorders of the urinary system |
| 600 | hyperplasia of prostate |
| 605 | erectile dysfunction |
| 608 | other disorders of male genital organs |
| 611 | abnormal findings on mammogram or breast exam |
| 612 | breast conditions, congenital or relating to hormones |
| 618 | genital prolapse |
| 622 | polyp of female genital organs |
| 623 | hypertrophy of female genital organs |
| 625 | pain and other symptoms associated with female genital organs |
| 626 | disorders of menstruation and abnormal bleeding from female genital tract |
| 627 | menopausal and postmenopausal disorders |
| 722 | intervertebral disc disorders |
| 731 | osteitis deformans and osteopathies associated with other disorders |
| 735 | acquired foot deformities |
| 740 | osteoarthrosis |
| 751 | genitourinary congenital anomalies |
| 755 | congenital anomalies of limbs |
| 761 | cervicalgia |
| 771 | musculoskeletal symptoms referable to limbs |
| 773 | pain in limb |
| 800 | fracture of lower limb |
| 801 | fracture of ankle and foot |
| 805 | fracture of vertebral column without mention of spinal cord injury |
| 807 | fracture of ribs |
| 809 | fracture of unspecified bones |
| 830 | dislocation |
| 840 | sprains and strains |
| 859 | complication due to other implant or internal device |
| 872 | traumatic imputation |
| 916 | contusion |
| 938 | dermatitis due to solar radiation |
| 952 | spinal cord injury without evidence of spinal bone injury |
| 961 | poisoning by other anti-infectives |
| 977 | personal history of allergy to medicinal agents |
| 1008 | crushing or internal injury to organs |
| 1009 | injury, NOS |
| 1010 | other tests |
| 1011 | complications of surgical and medical procedures |
| 1089 | acquired absence of limb |

### Table EII - Cohort Demographics

|  | **All** | **Age < 26 y** | **Age 26-64 y** | **Age > 64 y** |
| --- | --- | --- | --- | --- |
| **N** | 139 | 7 | 54 | 78 |
| **Mean Age in years (SD)** | 62.5 (17.8) | 20.1 (4.3) | 49.5 (10.8) | 75.3 (6.3) |
| **Mean EHR Length in days Prior to Dx (SD)** | 978.1 (833.0) | 1031.6 (998.8) | 939.7 (882.9) | 999.9 (778.1) |
| **Mean Min IgG level Prior to diagnosis (mg/dL)** | 424.9 (130.4) | 525.5 (0.0) | 399.1 (141.3) | 433.9 (124.2) |
| **Percent diagnosed by Allergy / Immunology** | 22% | 14% | 20% | 23% |

### Figure E1 - Cohort Definition Flowchart

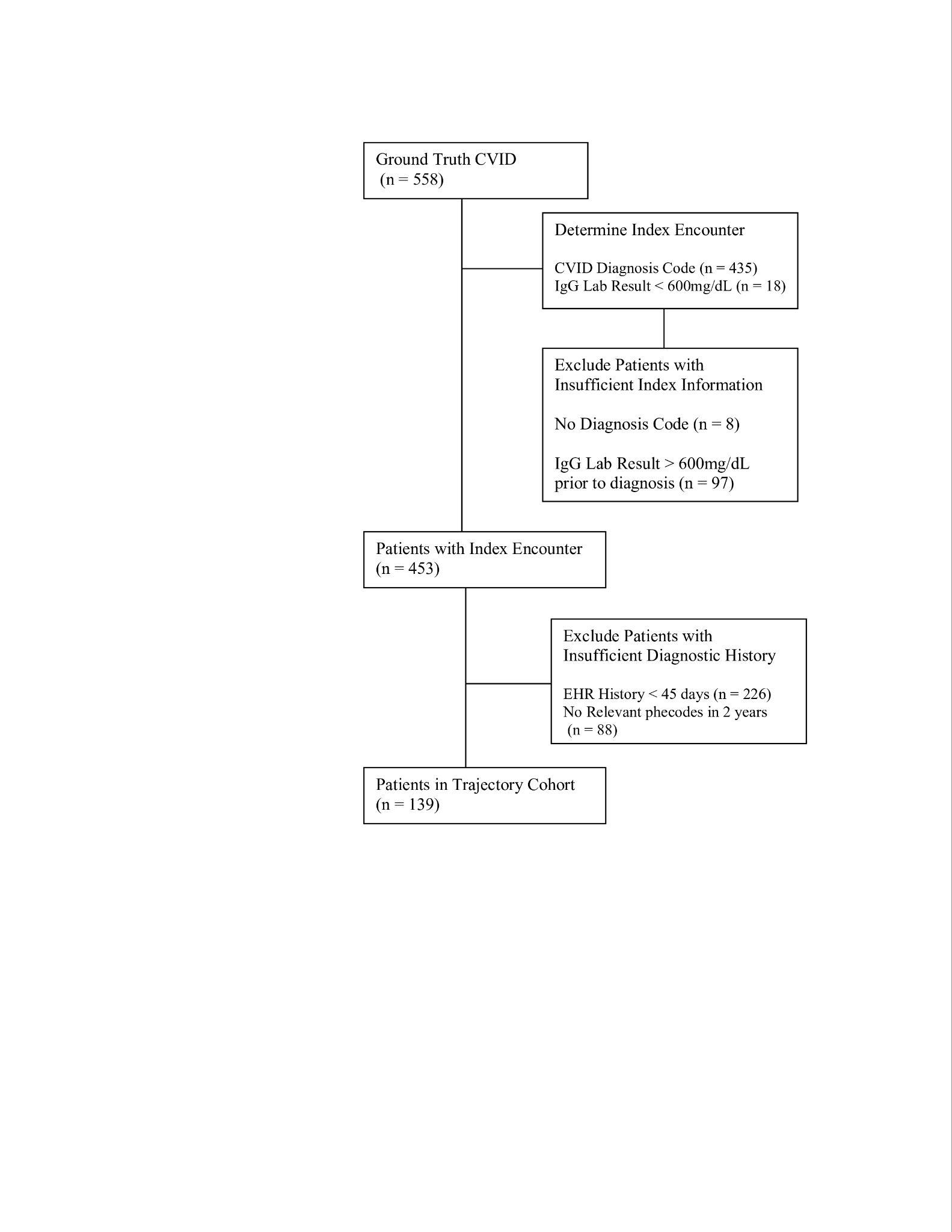

*Figure E1:* CVID patients go through rigorous quality control for inclusion in our cohort. Patient index encounters are defined based on a combination of ICD code and IgG lab result conditions, and patients with not enough documented clinical history or unclear index date were excluded from analysis.

### Figure E2 - Patient phecodes at CVID diagnosis

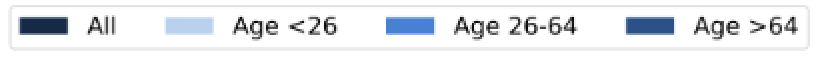

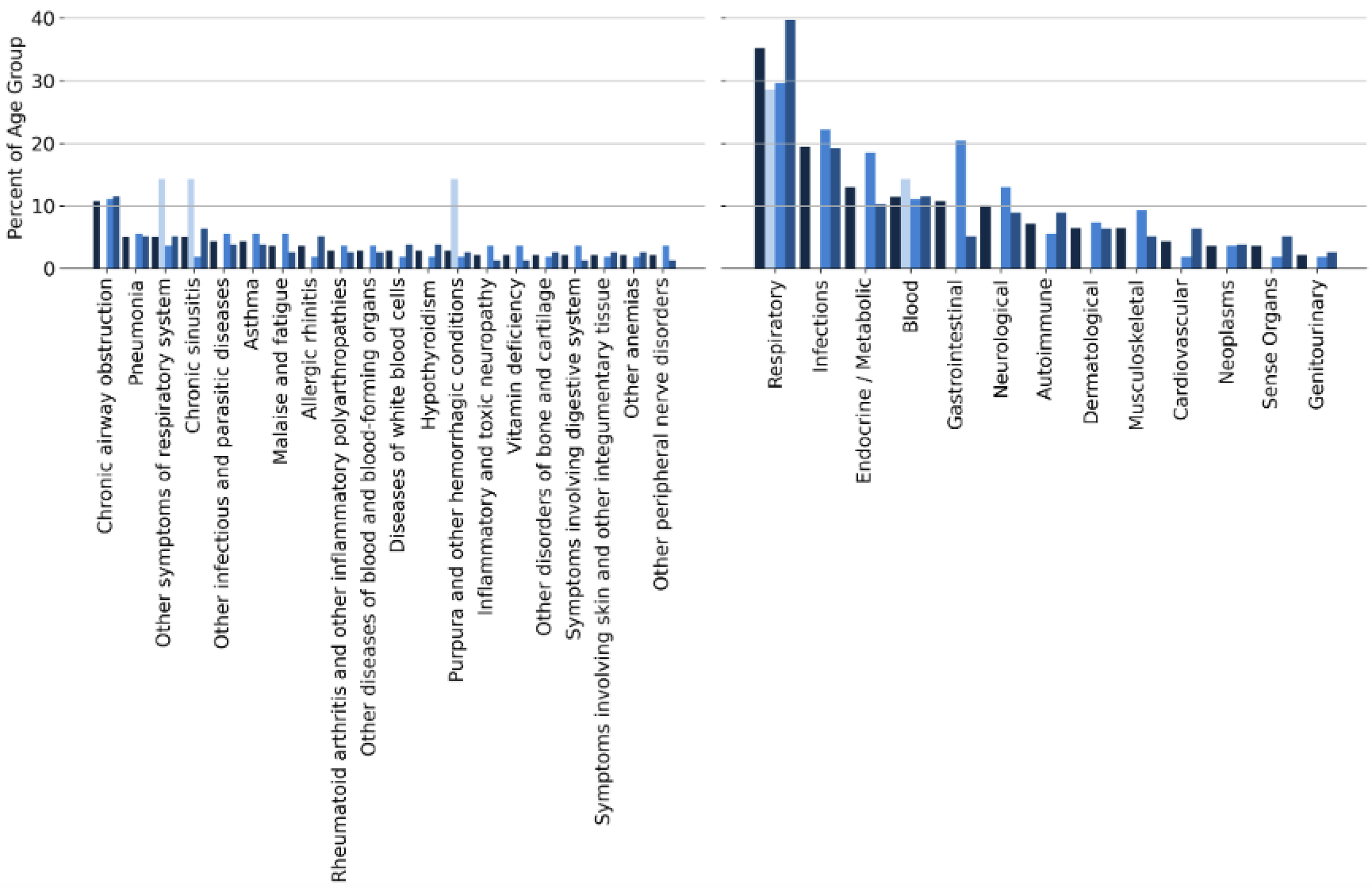

*Figure E2:* This figure shows patient symptoms at their CVID diagnosis visit (x axis). Colors differentiate between age groups. The left plot displays the top 20 phecodes present in our cohort at the diagnosis visit. The right plot displays these phecodes grouped by category.

### Figure E3 - Patient visit volume by specialty prior to CVID diagnosis

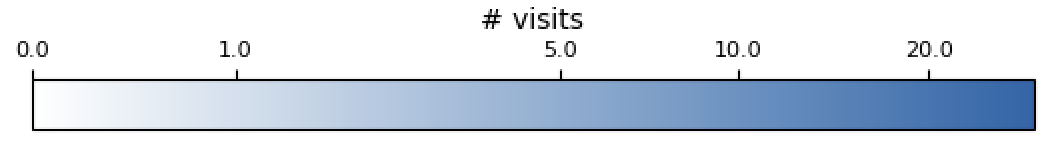

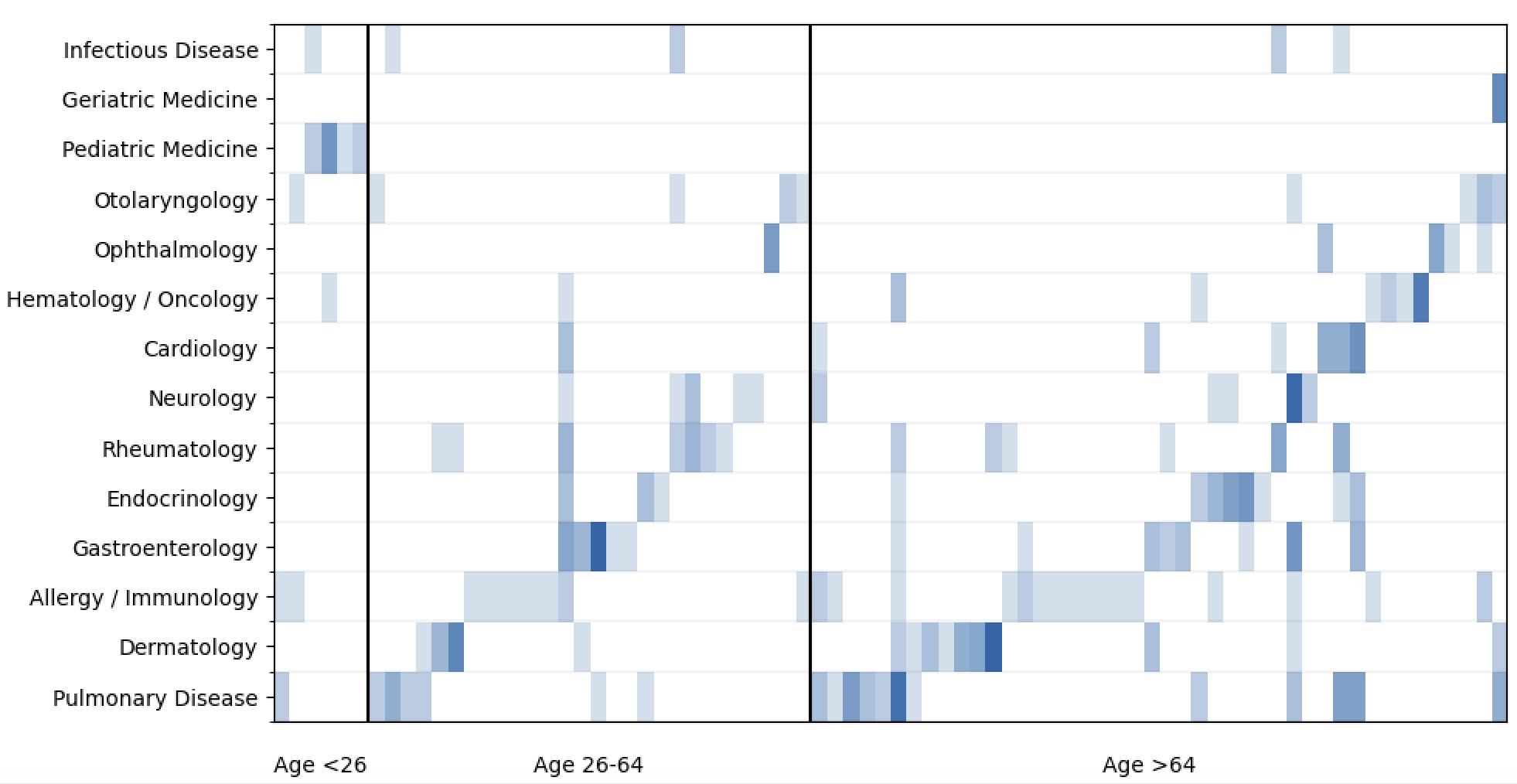

### *Figure E3:* Heatmap of CVID patient visits to various specialties in the two years prior to diagnosis ordered by patient age at diagnosis. Each specialty is a row and each patient is a column (patients are unlabeled to preserve unidentifiability). The heatmap is colored according to a log_2_(1+x) scale, with labels converted back to strict counts for readability. Average patients have three visits per year across 1.8 specialties.

### Figure E4: Distribution of visit history compared to matched controls

(a) Visits to Diagnosing Specialty	(b) Total Number of Specialties
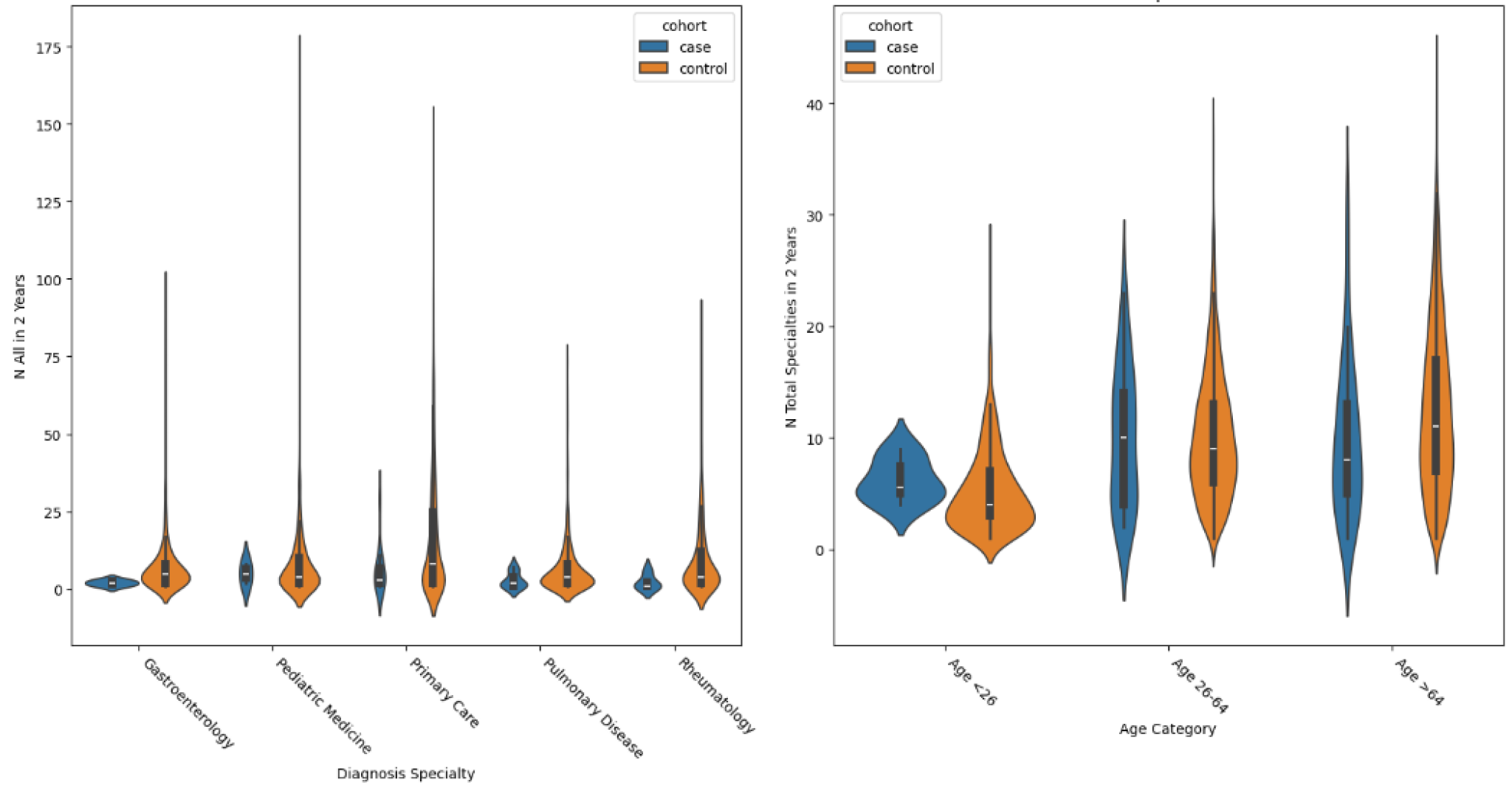

*Figure E4:* CVID patients (blue) have a different distribution of visits to their diagnosis specialty compared to other patients of those specialties (orange). The violin plot shows the median as well as the distribution for each column. Left: cases (blue) and matched controls (orange) number of visits (y-axis) to each specialty (x-axis) in the two years prior to CVID diagnosis (for cases) and in a two year time period (controls). Right: the total number of specialties (y-axis) seen in those two years for patients by age-group (x-axis).

### Figure E5 - Time-varying effects of specific symptoms on time to CVID diagnosis

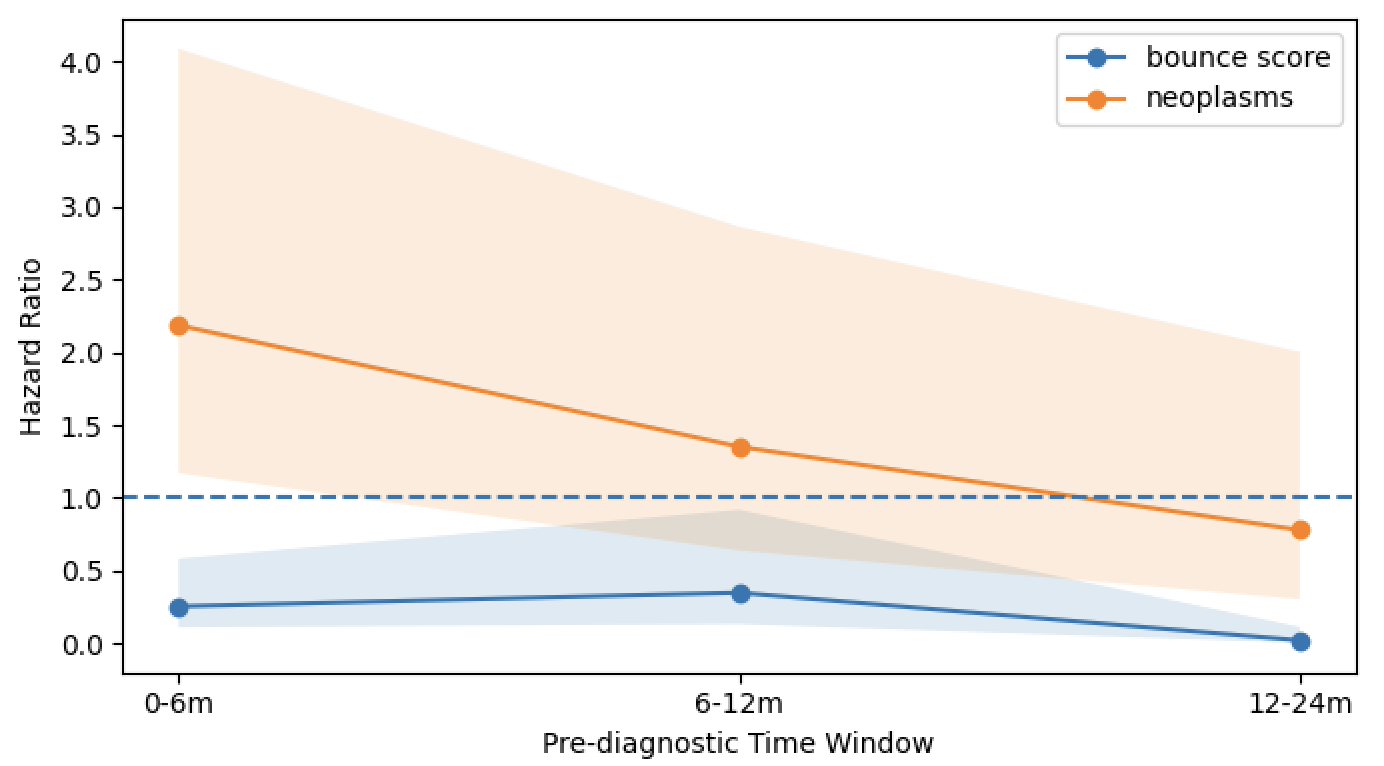

*Figure E5:* This figure shows the hazard ratio (y-axis) for neoplasms (orange) and bounce score (blue) as the pre-diagnostic time window (x-axis) varies. Features that remain on one side of the dotted line represent time-invariant features, whereas features that cross the dotted line represent time-specific features.

### Figure E6 - Key features driving specialty-specific CVID prediction

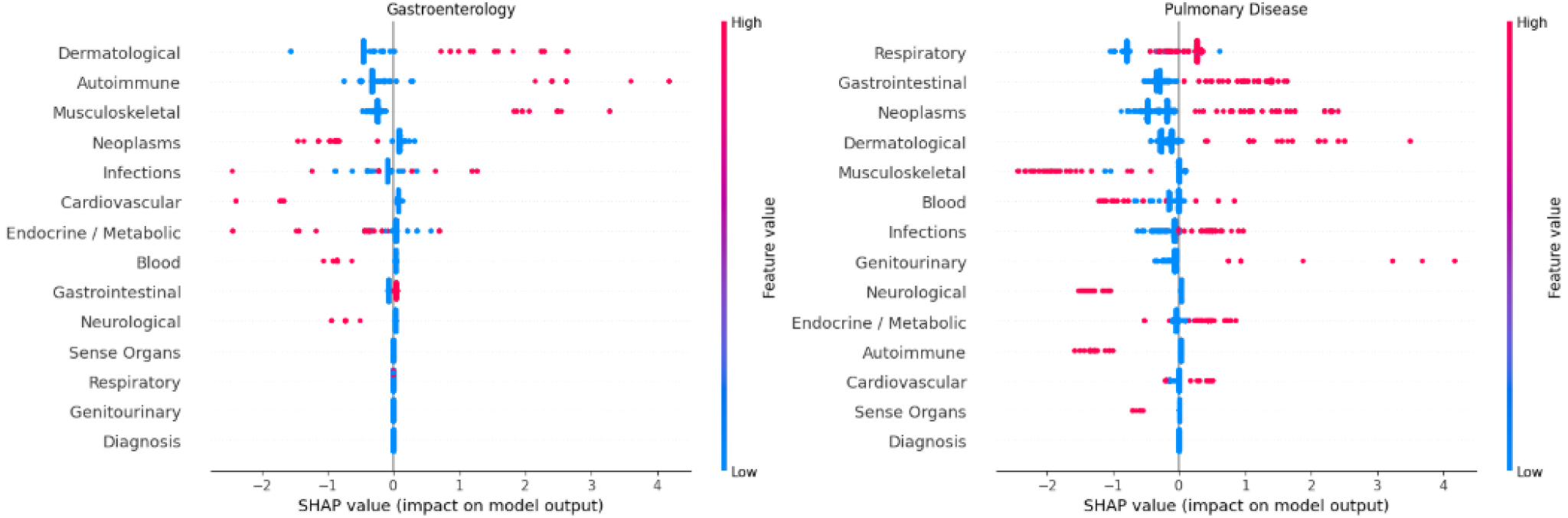

### *Figure E6:* This figure shows the Shapley Additive Explanations (SHAP) values for the specialty-specific XGBoost model predicting CVID. Each panel represents a different specialty, and within each panel features are ordered by overall importance. The color of each point represents the relative feature value for the model, the horizontal position indicates the direction of effect of the feature at that value on the model, and the vertical width the magnitude of effect. Positive values indicate a higher probability of CVID.

### Figure E7 - XGBoost Model performance of specialty-specific CVID prediction

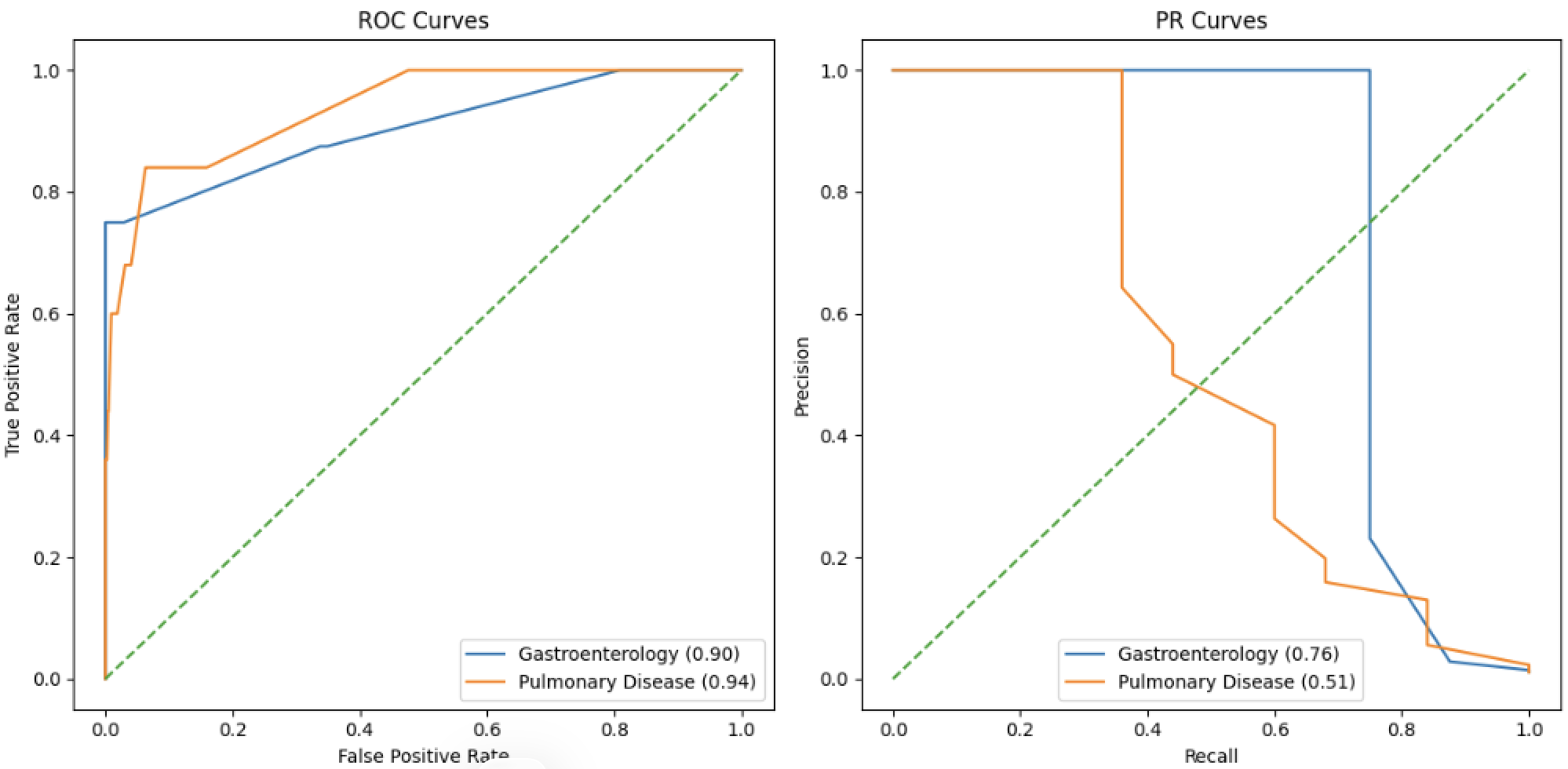

*Figure E7:* The left panel shows the receiver operator curve (ROC) for the XGBoost CVID prediction model, while the right panel shows the precision recall (PR) curve for the same model. XGBoost was run separately for each different specialty, shown in different colors.
